# Final sizes and durations of new COVID-19 pandemic waves in Poland and Germany predicted by generalized SIR model

**DOI:** 10.1101/2021.12.14.21267771

**Authors:** Igor Nesteruk

## Abstract

New waves of the COVID-19 pandemic in Europe, which began in the autumn of 2021, are a matter of great concern and the need to immediately predict the epidemic dynamics in order to assess the possible maximum values of new cases, the risk of infection and the number of deaths. The generalized SIR-model and corresponding parameter identification procedure was used to simulate and predict the dynamics of new epidemic waves in Poland and Germany. Results of calculations show that new cases in these countries will not stop to appear in 2022.

## Introduction

The COVID-19 pandemic dynamics in Poland was discussed in [1-9]. The pandemic spreading in Germany was investigated in [10-22]. In particular, to predict the first pandemic wave in Germany, the classical SIR model [23-25] and the statistics-based method of its parameter identification [26] were successfully used in [10, 14]. In the second SIR simulation for Germany [10, 14], the data set about the accumulated numbers of COVID-19 cases for the period: April 9–29, 2020 (according to WHO daily reports, [27]) was used. It was predicted that the first epidemic wave in Germany could stop on August 4, 2020 with the number of cases 176,952. The real accumulated number of cases was 212,022 (see [27]). It means that the accuracy of prediction was 16.5% after 95 days of pandemic monitoring (paper [10] was published in May 2020).

Changes in quarantine restrictions, social behavior, testing and vaccination levels, appearance of new strains, etc. cause the changes in the epidemic dynamics, i.e., so known new waves. To simulate new epidemic waves, a numerical method of their detection [28] was developed. In particular, thirteen epidemic waves were simulated for Ukraine and six pandemic waves for the whole world [14, 31, 34] with the use of a generalized SIR-model [29], and corresponding parameter identification procedures [30]. The correct selection of all the COVID-19 pandemic waves in Poland and Germany is still ahead. Here we will consider the new waves which occurred in October-December 2021 and predict their durations and final sizes with the use of generalized SIR-model.

### Data

We will use the data sets regarding the accumulated numbers of laboratory-confirmed COVID-19 cases in Poland and Germany reported by COVID-19 Data Repository by the Center for Systems Science and Engineering (CSSE) at Johns Hopkins University (JHU), [32]. The corresponding numbers *V*_*j j*_ and moments of time *t*_*j*_ (measured in days) are shown in Tables 1 and 2 for the period of July to December 2021. Here we use the datasets, corresponding to the period of November 22 to December 5, 2021 to simulate the 4th wave in Poland and the 5-th wave in Germany. Other *V*_*j*_ and *t*_*j*_ values will be used to control the accuracy of predictions and the pandemic dynamics.

**Table 1.** Cumulative numbers of laboratory-confirmed Covid-19 cases in Poland in the summer and autumn of 2021 according to the JHU datasets, [32].

| Day in corresponding month of 2021 | Number of cases in July, $V_j$ | Number of cases in August, $V_j$ | Number of cases in September $V_j$ | Number of cases in October, $V_j$ | Number of cases in November, $V_j$ | Number of cases in December, $V_j$ |
| --- | --- | --- | --- | --- | --- | --- |
| 1 | 2880010 | 2883029 | 2889036 | 2908432 | 3030151 | 3569137 |
| 2 | 2880107 | 2883120 | 2889412 | 2909776 | 3034668 | 3596491 |
| 3 | 2880215 | 2883284 | 2889773 | 2910866 | 3045102 | 3623452 |
| 4 | 2880270 | 2883448 | 2890161 | 2911549 | 3060613 | 3649027 |
| 5 | 2880308 | 2883624 | 2890484 | 2912876 | 3076518 | 3671421 |
| 6 | 2880403 | 2883796 | 2890666 | 2914962 | 3091713 | 3684671 |
| 7 | 2880503 | 2883976 | 2891071 | 2916969 | 3104220 | 3704040 |
| 8 | 2880596 | 2884098 | 2891602 | 2918863 | 3111534 | 3732589 |
| 9 | 2880670 | 2884162 | 2892113 | 2920874 | 3125179 | 3760048 |
| 10 | 2880755 | 2884361 | 2892642 | 2922401 | 3143725 | 3785036 |
| 11 | 2880821 | 2884557 | 2893173 | 2923304 | 3162804 | 3808798 |
| 12 | 2880865 | 2884780 | 2893649 | 2925425 | 3175769 |  |
| 13 | 2880959 | 2884974 | 2893919 | 2928065 | 3190067 |  |
| 14 | 2881046 | 2885185 | 2894455 | 2931064 | 3204515 |  |
| 15 | 2881151 | 2885333 | 2895223 | 2933834 | 3214023 |  |
| 16 | 2881241 | 2885461 | 2895947 | 2937069 | 3230634 |  |
| 17 | 2881355 | 2885676 | 2896599 | 2939590 | 3254875 |  |
| 18 | 2881424 | 2885883 | 2897395 | 2941126 | 3279787 |  |
| 19 | 2881491 | 2886079 | 2897935 | 2945056 | 3303046 |  |
| 20 | 2881594 | 2886291 | 2898299 | 2950616 | 3326464 |  |
| 21 | 2881718 | 2886513 | 2899008 | 2956207 | 3345388 |  |
| 22 | 2881840 | 2886698 | 2899888 | 2961923 | 3357763 |  |
| 23 | 2881948 | 2886805 | 2900862 | 2968200 | 3377698 |  |
| 24 | 2882066 | 2887037 | 2901674 | 2972927 | 3406129 |  |
| 25 | 2882146 | 2887270 | 2902591 | 2975880 | 3434272 |  |
| 26 | 2882220 | 2887485 | 2903234 | 2982143 | 3461066 |  |
| 27 | 2882327 | 2887739 | 2903655 | 2990509 | 3487254 |  |
| 28 | 2882465 | 2888028 | 2904631 | 2998891 | 3507828 |  |
| 29 | 2882630 | 2888231 | 2905866 | 3008294 | 3520961 |  |
| 30 | 2882786 | 2888385 | 2907071 | 3018100 | 3540061 |  |
| 31 | 2882939 | 2888670 | - | 3025247 | - |  |

**Table 2.** Cumulative numbers of laboratory-confirmed Covid-19 cases in Germany in the summer and autumn of 2021 according to the JHU datasets, [32].

| Day in corresponding month of 2021 | Number of cases in July, $V_j$ | Number of cases in August, $V_j$ | Number of cases in September, $V_j$ | Number of cases in October, $V_j$ | Number of cases in November, $V_j$ | Number of cases in December, $V_j$ |
| --- | --- | --- | --- | --- | --- | --- |
| 1 | 3736959 | 3778277 | 3979839 | 4249061 | 4619273 | 5999020 |
| 2 | 3737630 | 3779797 | 3993789 | 4255543 | 4649964 | 6072299 |
| 3 | 3738059 | 3782344 | 3996688 | 4260494 | 4684462 | 6134492 |
| 4 | 3738470 | 3786003 | 4005632 | 4265001 | 4722102 | 6177992 |
| 5 | 3738862 | 3789460 | 4013808 | 4272764 | 4755887 | 6200937 |
| 6 | 3739575 | 3792848 | 4020587 | 4284400 | 4779675 | 6219259 |
| 7 | 3740567 | 3795609 | 4039667 | 4295876 | 4792463 | 6312346 |
| 8 | 3741470 | 3797849 | 4044777 | 4305634 | 4816177 | 6339828 |
| 9 | 3742355 | 3800069 | 4068495 | 4312528 | 4857463 | 6442846 |
| 10 | 3743164 | 3803351 | 4071643 | 4318437 | 4908540 | 6496142 |
| 11 | 3743732 | 3808838 | 4080180 | 4323346 | 4957374 | 6528894 |
| 12 | 3744285 | 3814335 | 4087125 | 4331274 | 5002730 |  |
| 13 | 3745312 | 3819871 | 4093412 | 4343591 | 5037039 |  |
| 14 | 3746935 | 3824546 | 4102252 | 4355169 | 5056242 |  |
| 15 | 3748379 | 3828278 | 4115342 | 4366833 | 5091200 |  |
| 16 | 3749944 | 3831827 | 4127158 | 4375253 | 5144827 |  |
| 17 | 3751253 | 3837218 | 4137062 | 4382019 | 5213193 |  |
| 18 | 3752236 | 3846226 | 4144165 | 4388581 | 5271961 |  |
| 19 | 3753220 | 3854529 | 4149832 | 4410921 | 5329263 |  |
| 20 | 3754846 | 3863495 | 4155160 | 4429723 | 5374446 |  |
| 21 | 3756497 | 3870095 | 4162437 | 4447231 | 5400687 |  |
| 22 | 3758425 | 3876041 | 4173357 | 4462324 | 5448574 |  |
| 23 | 3760291 | 3881633 | 4183672 | 4466173 | 5516623 |  |
| 24 | 3761869 | 3898287 | 4192606 | 4476078 | 5595674 |  |
| 25 | 3763018 | 3911562 | 4199029 | 4485437 | 5670253 |  |
| 26 | 3764441 | 3923250 | 4204300 | 4516839 | 5735837 |  |
| 27 | 3766501 | 3925190 | 4209098 | 4545665 | 5780814 |  |
| 28 | 3769552 | 3933585 | 4216507 | 4553744 | 5804139 |  |
| 29 | 3772326 | 3940212 | 4228774 | 4591264 | 5854884 |  |
| 30 | 3774918 | 3947035 | 4239773 | 4608512 | 5923564 |  |
| 31 | 3776724 | 3965681 | - | 4607958 | - |  |

### Generalized SIR model and data smoothing procedure

The generalized SIR-model relates the number of susceptible *S*, infectious *I* and removed persons *R* for a particular epidemic wave *i*, [14, 29]. The exact solution of the set of non-linear differential equations uses the function

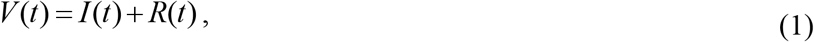

corresponding to the number of victims or the cumulative laboratory-confirmed number of cases [14, 29]. Its derivative:

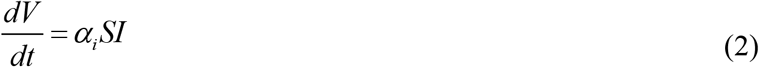

yields the estimation of the average daily number of new cases. When the registered number of victims *V*_*j*_ is a random realization of its theoretical dependence (1), the exact solution presented in [14, 29] depends on five parameters (*α*_*i*_ is one of them). The details of the optimization procedure for their identification can be found in [30].

Since daily numbers of new cases are random and characterized by some weekly periodicity, we will use the smoothed daily number of accumulated cases:

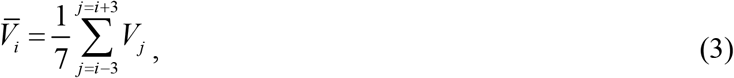

and its numerical derivative:

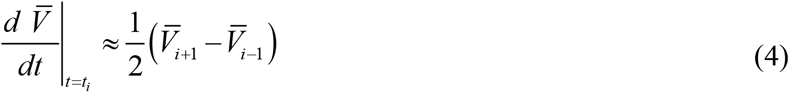

to estimate the smoothed number of new daily cases [10, 14, 28].

## Results and discussion

The optimal values of the general SIR model and other characteristics of the 4th epidemic wave for Poland and the 5th one in Germany are calculated and listed in Table 3. For comparison, the results for the 13th pandemic wave in Ukraine (calculated in [31]) are also presented in the last column. The corresponding SIR curves are shown in the figure by red lines for Poland and black lines for Germany. The pandemic dynamics in both countries is rather irregular. In particular, the smoothed numbers of new daily cases (4) have multiple minima after August 15, 2021 (see red and black “crosses” in the figure). It means that pandemic dynamics changed very rapid and many short epidemic waves occurred. In particular, a distinct epidemic wave in Germany in August – September 2021 is visible (see black “crosses”). That is why we use different numbers for autumn epidemic waves in Poland and Germany.

**Table 3.**
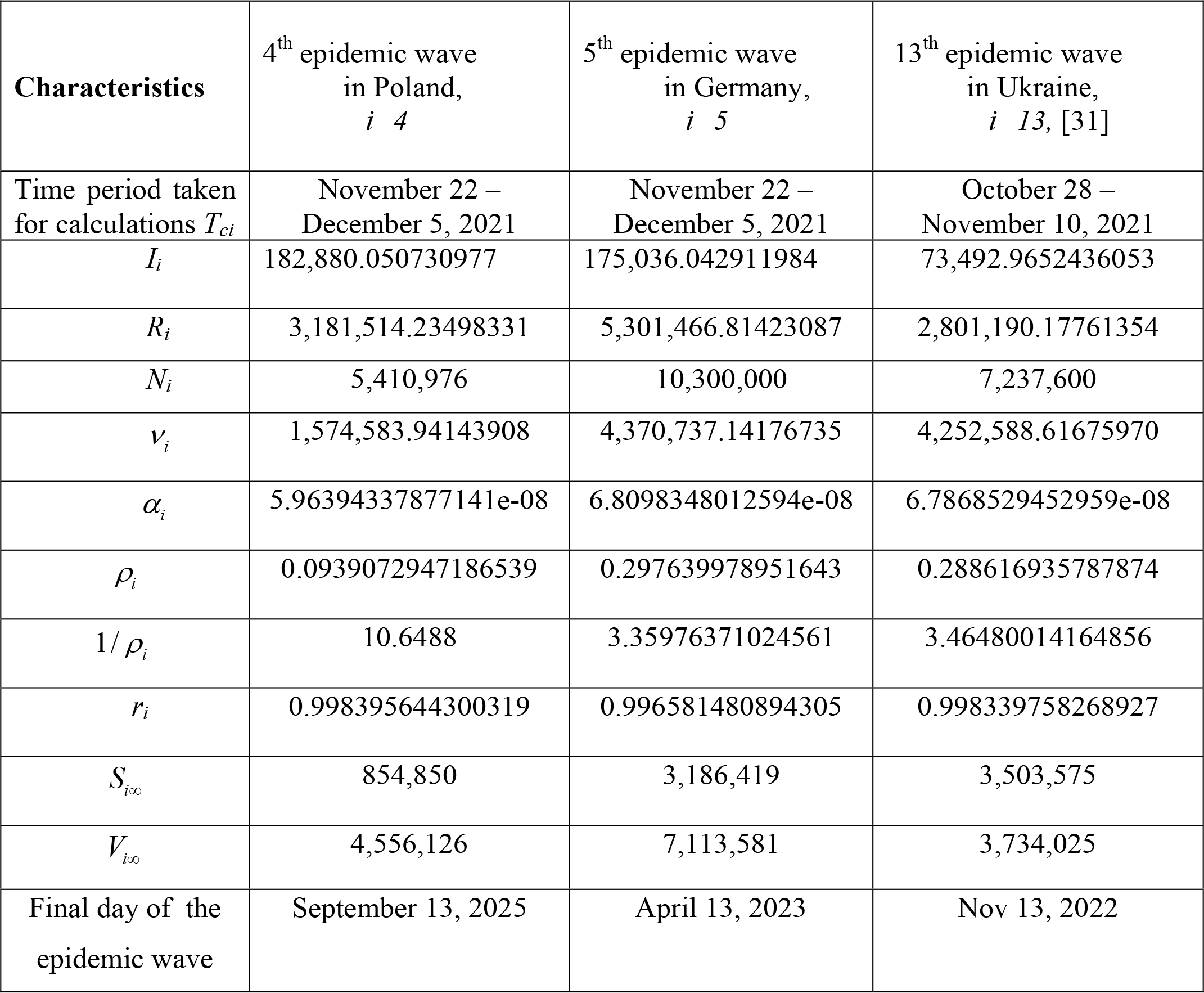
Optimal values of parameters and other characteristics the COVID-19 pandemic waves in Poland, Germany and Ukraine.

For correct SIR simulations, we need to separate periods with more or less stable dynamics first (see [14, 34]). Unfortunately, for the period taken for SIR simulations of pandemic waves in Poland and Germany (November 22 – December 5, 2021), some local minima of the smoothed numbers of new daily cases (4) are visible (see red and black “crosses” in the figure). This fact reduces the accuracy of SIR simulations and predictions. In particular, some problems with the identifications of the optimal values of model parameters occurred.

Table 3 demonstrates that the optimal values of SIR parameters are very different. Close values were obtained only for the average times of spreading the infection 1/ *ρ*_*i*_ for Germany and Ukraine. The assessments of the pandemic wave durations (corresponding the moment when the number of infectious persons becomes less that unit) are very pessimistic (April, 2023 for Germany and September 2025 for Poland). Similar long epidemic waves were predicted for India [33] and the whole world [31] with the use of the same generalized SIR model. In particular, if the global situation with vaccination, testing and treatment will not change, the COVID-19 pandemic could continue for another ten years [31].

**Figure.**
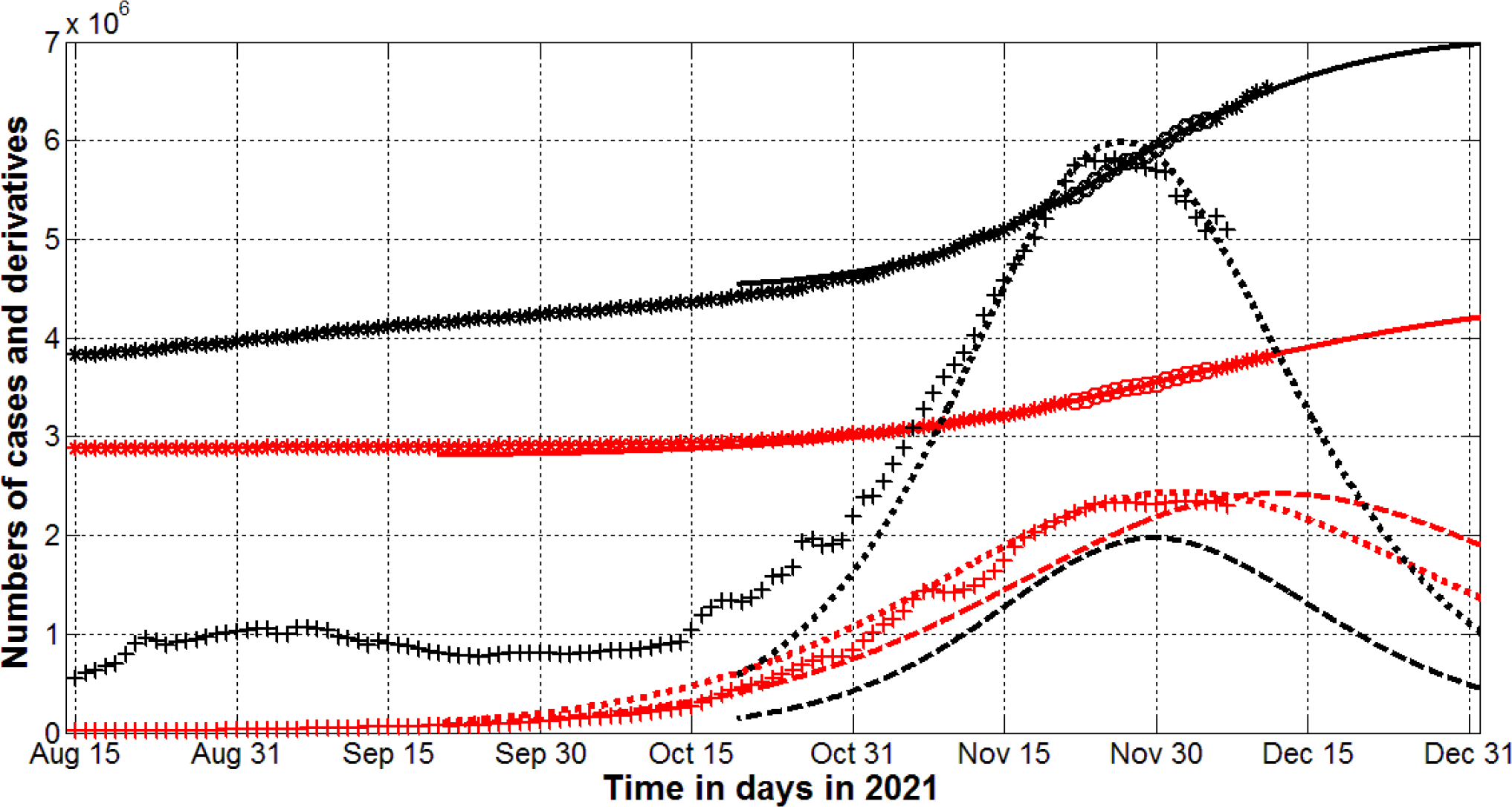
The COVID-19 pandemic waves in Poland (red) and Germany (black) in the autumn of 2021. The results of SIR simulations of the 4th wave in Poland and the 5th wave in Germany are shown by red and black lines, respectively. Numbers of victims *V(t)=I(t)+R(t)* – solid lines; numbers of infected and spreading *I(t)* multiplied by 10 – dashed; derivatives *dV/dt* (eq. (2), multiplied by 100) – dotted. “Circles” correspond to the accumulated numbers of cases registered during the period of time taken for SIR simulations. “Stars” correspond to *V*_*j*_ values beyond this time period. “Crosses” show the first derivative (4) multiplied by 100.

“Stars” and “crosses” in the figure illustrate the accuracy of simulations for the accumulated number of cases and the averaged daily numbers of new cases (eq. (4)). Comparisons with corresponding solid and dotted lines show that the theoretical estimations are consistent with observations for Poland. In the case of Germany the theoretical simulation are close to observations after Nobember 1, 2021. The theory predicts the decrease of the daily numbers of new cases after December 1, 2021 in both countries (see black and red dotted lines). The rate of this decreasing will be higher in Germany and by the end of 2021, the daily number of new cases in Germany is expected to be lower than in Poland. A significant increase in contacts during the New Year and Christmas holidays or/and the wide spread of new coronavirus strains (e.g., omicron) could disrupt these positive trends.

SIR simulations allows us to estimate the numbers of infectious persons *I(t)* and the probabilities to meet such a person in a population. Dashed lines show that the numbers of infectious in Poland are higher than in Germany and are expected to be much higher by the end of 2021. Taking into account the difference in populations of these countries, we can conclude, that contacts during the holidays in Poland are much more dangerous.

Unfortunately, the general SIR model cannot predict the emergence of new epidemic waves. It simulates the dynamics for only the period with constant epidemic conditions. Therefore, permanent monitoring of the number of new cases is needed to determine changes in the epidemic dynamics. After that it will be possible to perform new simulations by means of the generalized SIR model with calculation and use of new values of its parameters.

## Conclusions

The generalized SIR-model and corresponding parameter identification procedure was used to simulate and predict the dynamics of two new epidemic waves in Poland and Germany. If the situation with vaccination, testing and treatment will not change, the new COVID-19 will not stop to appear in 2022.

## Data Availability

All data produced in the present work are contained in the manuscript

## Acknowledgements

The author is grateful to Oleksii Rodionov for his help in collecting and processing data.

